# First cases of infection with the 21L/BA.2 Omicron variant in Marseille, France

**DOI:** 10.1101/2022.02.08.22270495

**Authors:** Philippe Colson, Jérémy Delerce, Mamadou Beye, Anthony Levasseur, Céline Boschi, Linda Houhamdi, Hervé Tissot-Dupont, Nouara Yahi, Matthieu Million, Bernard La Scola, Jacques Fantini, Didier Raoult, Pierre-Edouard Fournier

**Affiliations:** IHU Méditerranée Infection, 19-21 boulevard Jean Moulin, 13005 Marseille, France; Aix-Marseille Univ., Institut de Recherche pour le Développement (IRD), Microbes Evolution Phylogeny and Infections (MEPHI), 27 boulevard Jean Moulin, 13005 Marseille, France; Assistance Publique-Hôpitaux de Marseille (AP-HM), 264 rue Saint-Pierre, 13005 Marseille, France; Aix-Marseille Université, INSERM UMR S 1072, 51 boulevard Pierre Dramard, 13015 Marseille, France; Aix-Marseille Univ., Institut de Recherche pour le Développement (IRD), Vecteurs – Infections Tropicales et Méditerranéennes (VITROME), 27 boulevard Jean Moulin, 13005 Marseille, France

**Author notes:** **Corresponding authors:** Philippe Colson, IHU Méditerranée Infection, 19-21 boulevard Jean Moulin, 13005 Marseille, France.; Pierre-Edouard Fournier, IHU Méditerranée Infection, 19-21 boulevard Jean Moulin, 13005 Marseille, France.

**Keywords:** SARS-CoV-2, variant, Omicron, travel, emergence, southern France

## Abstract

The SARS-CoV-2 21K/BA.1, 21L/BA.2, and BA.3 Omicron variants have recently emerged worldwide. To date, the 21L/BA.2 Omicron variant has remained very minority globally but became predominant in Denmark instead of the 21K/BA.1 variant. Here we describe the first cases diagnosed with this variant in south-eastern France. We identified thirteen cases using variant-specific qPCR and next-generation sequencing between 28/11/2021 and 31/01/2022, the first two cases being diagnosed in travellers returning from Tanzania. Overall, viral genomes displayed a mean (±standard deviation) number of 65.9±2.5 (range, 61-69) nucleotide substitutions and 31.0±8.3 (27-50) nucleotide deletions, resulting in 49.6±2.2 (45-52) amino acid substitutions (including 28 in the spike protein) and 12.4±1.1 (12-15) amino acid deletions. Phylogeny showed the distribution in three different clusters of these genomes, which were most closely related to genomes from England and South Africa, from Singapore and Nepal, or from France and Denmark. Structural predictions pointed out a significant enlargement and flattening of the 21L/BA.2 N-terminal domain surface compared with that of the 21K/BA.2 Omicron variant, which may facilitate initial viral interactions with lipid rafts. Close surveillance is needed at global, country and center scales to monitor the incidence and clinical outcome of the 21L/BA.2 Omicron variant.

## Introduction

SARS-CoV-2 variants have been detected since summer 2020^1,2^ and revealed of critical interest regarding viral transmissibility, load, and escape to natural or vaccine immunity.^3,4^ The Omicron variant is currently the predominant variant of concern in many countries worldwide (https://covariants.org/per-country).^5,6^ It has been reported to exhibit considerable escape to antibodies elicited by vaccination^7,8^ and to be associated with lower clinical severity including in our center.^8,9^ It was first detected in early November in Botswana and thereafter, in many countries, its incidence has rapidly exceeded that of the Delta variant that had predominated since the summer of 2021 (https://covariants.org/per-country).^5,6^ As a matter of fact, Omicron is composed of three branches corresponding to three variants named Nextstrain clade^10,11^ 21K (or Pangolin lineage^12^ BA.1), 21L (or BA.2), and lineage BA.3 that is part of the 21M Omicron clade. Until very recently, unlike the 21K/BA.1 Omicron variant, the 21L/BA.2 Omicron variant has remained a very minority in most countries and globally, including in South Africa from where it seems to originate. Here we describe the emergence in south-eastern France of this variant.

## Materials and methods

Nasopharyngeal samples were collected from patients in our university hospital institute (Méditerranée Infection; https://www.mediterranee-infection.com/) and tested for SARS-CoV-2 infection by real-time reverse transcription PCR (qPCR) as previously described.^2,13^ Then qPCR assays specific of variants were performed according to French recommendation, as previously reported.^2,13,14^ This included detection of spike mutations L452R, K417N, E484K, and/or P681H (Thermo Fisher Scientific, Waltham, USA), combined with testing with the TaqPath COVID-19 kit (Thermo Fisher Scientific) that target viral genes ORF1, N (nucleocapsid) and S (spike).

Genomic identification of the 21L/BA.2 Omicron variant was performed through next-generation sequencing with the Oxford Nanopore technology (ONT) on a GridION instrument (Oxford Nanopore Technologies Ltd., Oxford, UK) or with the Illumina COVID-seq protocol on the NovaSeq 6000 instrument (Illumina Inc., San Diego, CA, USA), as previously described.^2,13,14^ Sequence read processing and genome analysis were performed as previously described.^2,13,14^ Fastq files were processed differently according to the sequencing technology. Briefly, for ONT reads, fastq files were processed with the ARTIC field bioinformatics pipeline (https://github.com/artic-network/fieldbioinformatics). Sequencing reads were basecalled with Guppy (v.4.0.14) and aligned to the Wuhan-Hu-1 genome GenBank accession no. NC_045512.2 using minimap2 (v2.17-r941) (https://github.com/lh3/minimap2). Reads were cleaned with Guppyplex. Mapping was cleaned with ARTIC align_trim. Variant calling was performed using Medaka and Longshot. Consensus genome sequences were built with Bcftools. Illumina NovaSeq reads were basecalled with the Dragen Bcl Convert pipeline (v3.9.3; https://emea.support.illumina.com/sequencing/sequencing_software/bcl-convert.html (Illumina Inc.)), mapping was performed with the bwa-mem2 tool (https://github.com/bwa-mem2/bwa-mem2) on the Wuhan-Hu-1 genome. Mapping was cleaned with Samtools (https://www.htslib.org/). Variant calling was performed with freebayes (https://github.com/freebayes/freebayes) and consensus genomes were built with Bcftools (https://samtools.github.io/bcftools/bcftools.html).

Nucleotide and of amino acid changes in viral genomes relatively to the Wuhan-Hu-1 isolate genome were obtained using the Nextclade tool (https://clades.nextstrain.org/).^10,11^ Nextstrain clades and Pangolin lineages were determined using the Nextclade web application (https://clades.nextstrain.org/)^10,11^ and Pangolin web application (https://cov-lineages.org/pangolin.html)^12^, respectively. Genome sequences described here were deposited in the GISAID sequence database (https://www.gisaid.org/) (Table 1).^15^ Finally, phylogeny was reconstructed with the nextstrain/ncov tool (https://github.com/nextstrain/ncov) then visualized with Auspice (https://docs.nextstrain.org/projects/auspice/en/stable/). The genomes the closest genetically to those obtained here were selected using Usher (https://genome.ucsc.edu/cgi-bin/hgPhyloPlace) and the GISAID BLAST tool (https://www.epicov.org/epi3/) then incorporated in phylogeny with all 21L/BA.2 Omicron variant genomes from France available in GISAID.

**Table 1.**
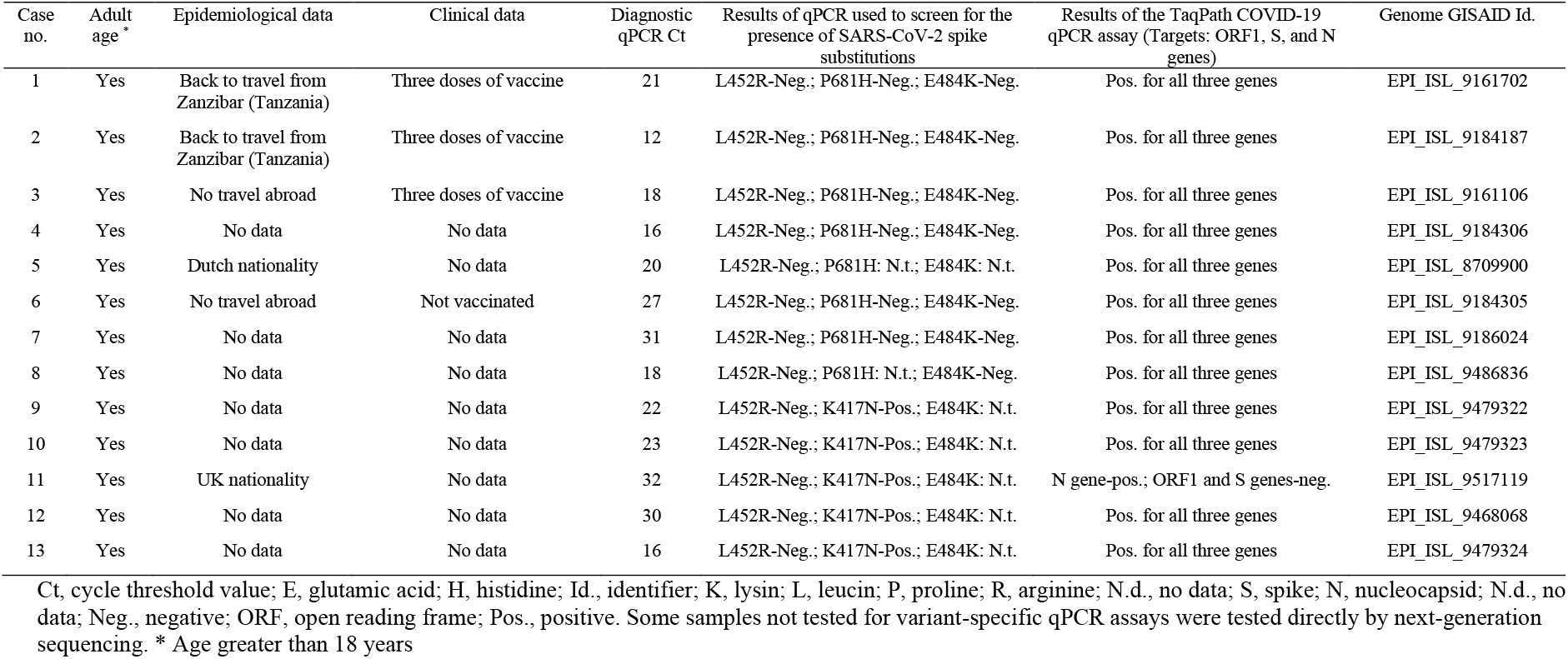
Main epidemiological and virological features of cases identified with infection with the SARS-CoV-2 21L/BA.2 Omicron variant.

This study was approved by the ethics committee of University Hospital Institute Méditerranée Infection (N°2022-008). Access to patients ‘ biological and registry data issued from the hospital information system was approved by the data protection committee of Assistance Publique-Hôpitaux de Marseille and recorded in the European General Data Protection Regulation registry under number RGPD/APHM 2019-73.

## Results

Thirteen infections with the 21L/BA.2 Omicron variant were diagnosed in our university hospital institute from patients sampled between 27/12/2021 and 31/01/2022 (Table 1). First cases were in two spouses in their 60s diagnosed late 2021 five days after returning from a travel in Zanzibar, Tanzania. They received a third dose of Pfizer-BioNTech COVID-19 vaccine three weeks before diagnosis. The third case had contacts with migrant patients of unknow SARS-CoV-2 status and a SARS-CoV-2-positive case (not tested in our institute) who met students from different countries. This third patient received a third dose of Pfizer-BioNTech COVID-19 vaccine seven weeks before diagnosis. Two other patients originate from the Netherlands and the United Kingdom. No information was available for the other eight patients.

All 21L/BA.2 Omicron variant-positive respiratory samples exhibited the same combination of spike mutations as screened by real-time qPCR: negativity for L452R, and, when performed, positivity for K417N and P681H and negativity for E484K and P681R (Table 1). In addition, the TaqPath COVID-19 kit (Thermo Fisher Scientific, Waltham, USA) provided positive signals for all three genes targeted (ORF1, S, and N), except for one sample that showed positivity for the N gene but negativity for both ORF1 and S genes, which was most likely due to a low viral load (qPCR cycle threshold, 32). Thus, 21L/BA.2 Omicron variant-infected patients could be distinguished by qPCR screening from the Delta (L452R-positive) and Omicron 21K (negative for S gene detection by the TaqPath COVID-19 assay) variants that co-circulated in southern France at the time of Omicron 21L/BA.2 emergence.

Thirteen 21L/BA.2 Omicron genomes were obtained. Analysis of those larger than 28,000 nucleotides showed the presence of a mean (±standard deviation) of 65.9±2.5 (range, 61-69) nucleotide substitutions and 31.0±8.3 (27-50) nucleotide deletions, which resulted in 49.6±2.2 (45-52) amino acid substitutions and 12.4±1.1 (12-15) amino acid deletions. All nine patients ‘ viruses harboured the same set of 28 amino acid substitutions and three contiguous amino acid deletions in their spike protein (Figure 1a). In addition, they shared the same set of amino acid substitutions: these included (i) seven substitutions located in other structural proteins (4, 2 and one in the nucleocapsid, membrane, and envelope proteins, respectively); (ii) 12 substitutions located in non-structural proteins including four in Nsp4, two in Nsp3 (a papain-like protease with phosphatase activity^16^, and one each in Nsp1, Nsp5 (a 3C-like proteinase), Nsp12 (RNA-dependent RNA polymerase), Nsp13 (helicase), Nsp14 (3 ’-5 ’-exonuclease with proofreading activity), and Nsp15 (an endoribonuclease); and (iii) one substitution located in ORF9b, a regulatory protein. Finally, three contiguous amino acid deletions were located in the nucleocapsid protein and three others were located in ORF9b. Of the 28 amino acid substitutions present in the spike of the 21L/BA.2 Omicron variant, 20 are shared with the 21K/BA.1 as well as the BA.3 Omicron variants (https://covariants.org/variants/) (Figure 1a).^5,6,17^

**Figure 1.**
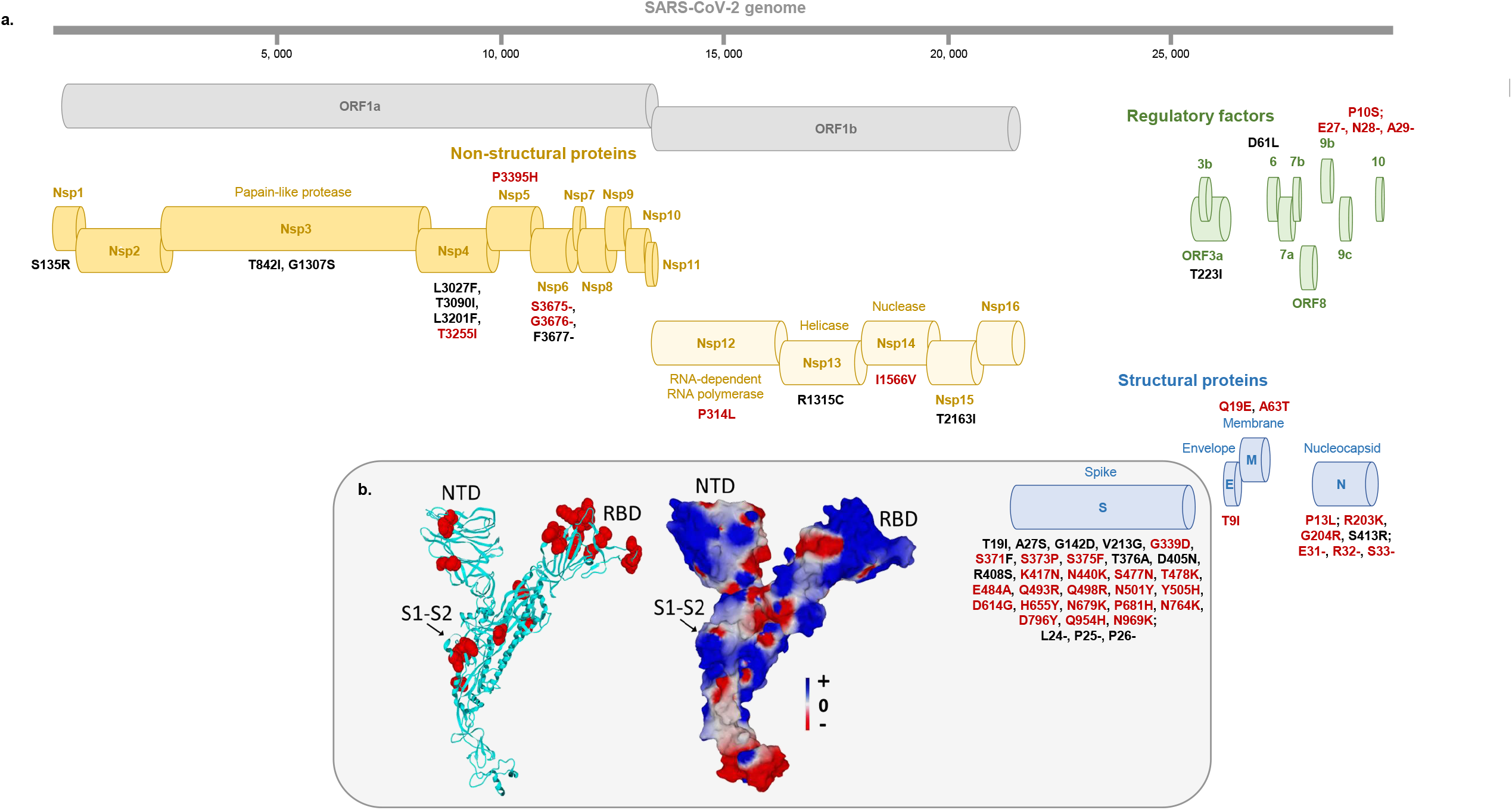
Map of the Omicron 21L/BA.2 spike protein with signature amino acid substitutions and deletions (a) and structural features of 21L/BA.2 Omicron variant spike protein (b) a: Amino acid substitutions and deletions shared with the 21K/BA.1 Omicron variant are indicated by a red font. b: Structural model of the Omicron 21L/BA.2 spike protein with mutations highlighted in red atomic spheres (left panel) or in electrostatic surface rendering (right panel). Note the flat surface of the N-terminal domain that faces lipid rafts of the host cell membrane. The S1-S2 cleavage site is indicated by an arrow. The color scale for the electrostatic surface potential (negative in red, positive in blue, neutral in white) is indicated. NTD, N-terminal domain; RBD, Receptor binding domain.

Phylogeny performed with the nextstrain/ncov tool (https://github.com/nextstrain/ncov) shows that the nine 21L/BA.2 Omicron variant genomes obtained in our institute were part of three clusters. Two genomes that were retrieved from the two patients who travelled in Tanzania were clustered with genomes obtained in England and South Africa (Figure 2). The genome retrieved from the Dutch patient was clustered with two genomes obtained in Nepal and Singapore. All other six genomes were most closely related to genomes from France and Denmark. As the first two cases we diagnosed were most likely infected with the 21L/BA.2 Omicron variant during their travel in Tanzania, we sought for this variant in GISAID among genomes from this country, but as of 02/02/2022 only three genomes (EPI_ISL_8917336, EPI_ISL_8917337, EPI_ISL_9391124) were available from this country: they were obtained from samples collected in December 2021 and belong to the 21K/BA.1 Omicron variant.

**Figure 2.**
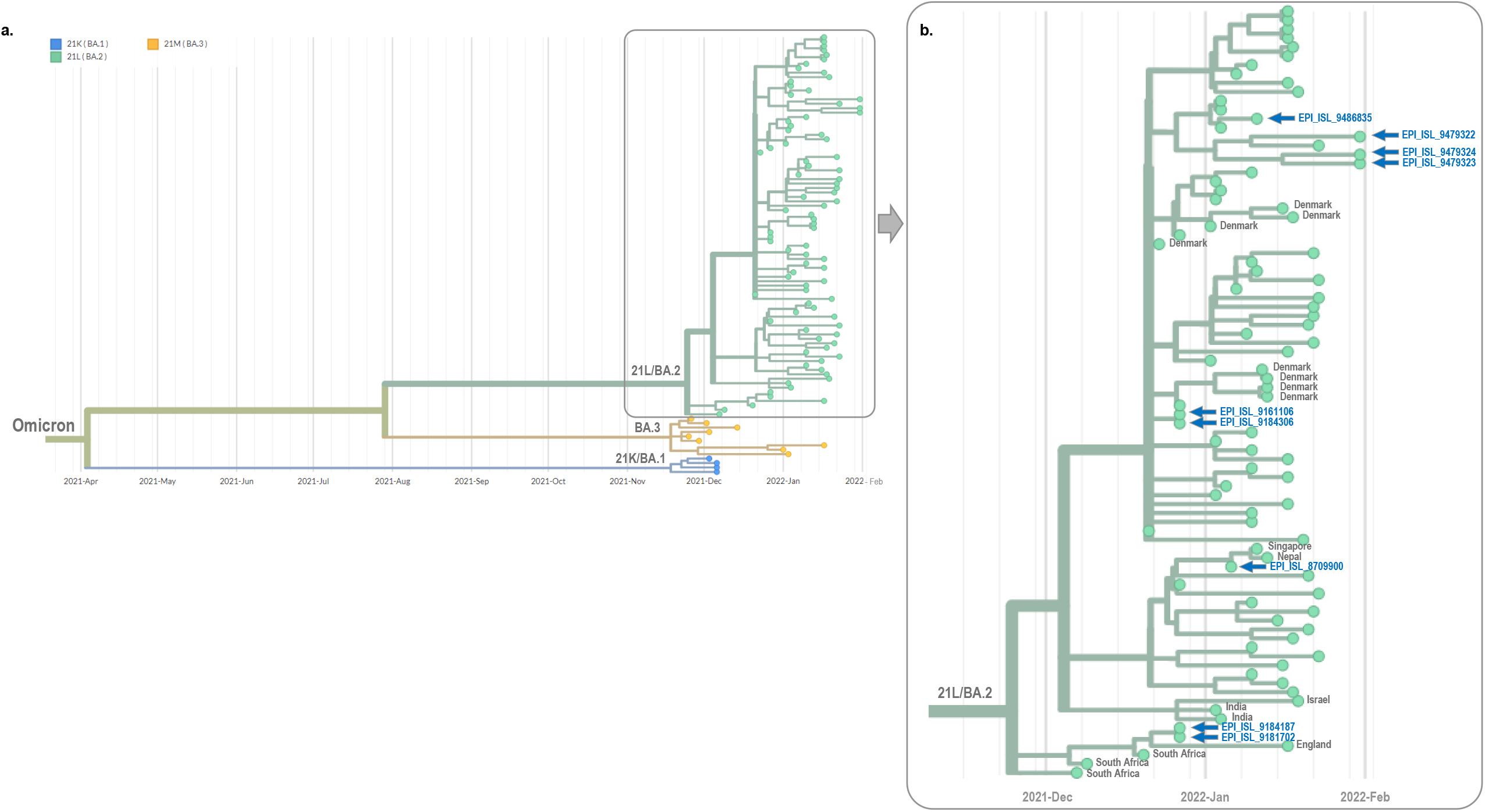
Phylogeny reconstruction based on genomes of the 21L/BA.2 Omicron variant obtained in the present study. Figure 2a incorporated genome sequences of 21K/BA.1, 21L/BA.2 and BA.3 Omicron variants. Figure 2b is a zoom of the 21L/BA.2 Omicron cluster of Figure 2a. Phylogenetic tree was built using the nextstrain/ncov tool (https://github.com/nextstrain/ncov) then visualized with Auspice (https://docs.nextstrain.org/projects/auspice/en/stable/). X-axis shows time. The 21L/BA.2 Omicron genomes the closest genetically to those obtained in our institute were selected using the Usher tool (https://genome.ucsc.edu/cgi-bin/hgPhyloPlace) and the GISAID BLAST tool (https://www.epicov.org/epi3/) and they were incorporated in the phylogenetic analysis in addition to all 21L/BA.2 Omicron variant genomes from France available in GISAID as of 02/02/2022. Sequences obtained in our laboratory (IHU Méditerranée Infection, Marseille, France) are indicated by a dark blue arrow and their GISAID identifier is indicated. Countries are indicated when they are not France.

The earliest 21L/BA.2 Omicron variant genome available from GISAID was obtained in South Africa from a sample collected on 17/11/2021 (EPI_ISL_6795834). As of 02/02/2022, most of the 37,521 21L/BA.2 Omicron variant genomes were obtained in Denmark (n= 24,138; 64%) (Figure 3a). Other countries with the greatest number of genomes were United Kingdom (n= 4,637 cases; 12%), India (n= 3,073 cases; 8%), Germany (n= 1,104 cases; 2.9%), and Philippines (n= 890 cases; 2.4%). Overall, Europe, Asia, North America, Africa and Oceania accounted for 34,498, 5,071, 398, 377, and 184 genomes, respectively. South Africa, where the 21L/BA.2 Omicron was first described, and Botswana accounted for only 304 and 62 genomes, respectively, amongst 5,550 and 1,449 genomes deposited in GISAID and obtained from samples collected since 01/12/2021, respectively. Finally, only 86 genomes (0.2%) were available for France out of 38,350 genomes deposited in GISAID and obtained from samples collected since 01/12/2021, while 18,219 21K/BA.1 Omicron variant genomes (48%) were available for the same period of time.

**Figure 3.**
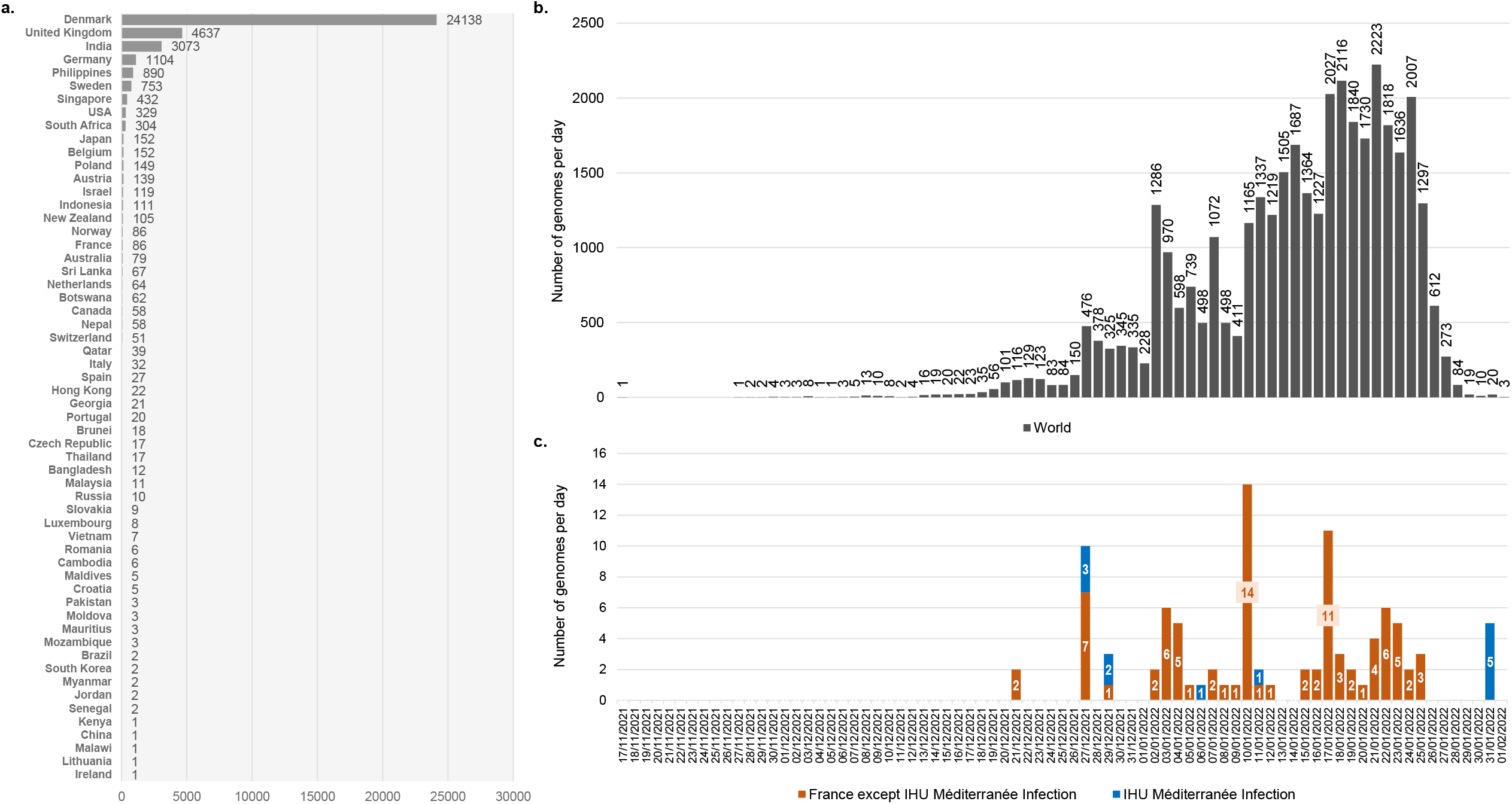
Number of genomes of the SARS-COV-2 21L/BA.2 Omicron variant available in GISAID and chronology of collections of respiratory samples from where they were obtained. a: Number of genomes of the SARS-COV-2 21L/BA.2 Omicron variant available in the GISAID sequence database (https://www.gisaid.org/)^15^ as of 02/02/2022. b: Chronology of SARS-CoV-2 diagnoses with the 21L/BA.2 Omicron variant for genomes deposited in the GISAID sequence database and obtained worldwide. c: Chronology of SARS-CoV-2 diagnoses with the 21L/BA.2 Omicron variant for genomes deposited in the GISAID sequence database and obtained in France or in our university hospital institute. The number of genomes was analyzed until 02/02/2022. Total number of genomes analyzed was 36,428. A total of 1,093 genomes were excluded as the date of sample collection was uncomplete (days or months were lacking).

Molecular modeling of Omicron 21L/BA.2 spike protein was performed as previously described^18^ by introducing the appropriate mutations and deletions in the framework of a complete 14-1,200 structure of the original 20B SARS-CoV-2 (Wuhan-Hu-1 isolate with D614G substitution)^18^ and by incorporating the missing amino acids with the Robetta protein structure prediction tool [https://robetta.bakerlab.org/] before energy minimization with the Polak-Ribière algorithm (Figure 1b).^19^ The new 21L/BA.2 Omicron variant displays several common structural features with its close relative, the 21K/BA.1 Omicron variant: many mutations exist that are chiefly distributed in the N-terminal domain (NTD), the receptor binding domain (RBD), and the S1-S2 cleavage site. As for the 21K/BA.1 Omicron variant, the electrostatic surface potential of the RBD is mostly positive, whereas the NTD is constituted by a patchwork of electronegative, electropositive, and neutral regions. A key difference between both 21L/BA.2 and 21K/BA.1 Omicron spike proteins is the significant enlargement and flattening of the 21L/BA.2 Omicron NTD surface compared with that of the 21K/BA.1 Omicron variant.^18^ This structural change is due to the lack of deletion 143-145 in the 21L/BA.2 Omicron variant. The flat surface of the 21L/BA.2 Omicron NTD may facilitate the initial interaction of the virus with lipid rafts,^19^ especially since the surface gain corresponds to an electropositive area (located on the left of the NTD in Figure 1b). Overall, one could hypothesize that the 21L/BA.2 Omicron variant NTD is better adapted to the electronegative surface of lipid rafts than that of the 21K/BA.1 Omicron variant.

## Discussion

It is currently unknown if this 21L/BA.2 Omicron variant would rise considerably in prevalence and compete the currently predominant Omicron 21K/BA.1, which has spread massively and quickly in countries with a high level of vaccine coverage.^5^ However, the very recent rise of the 21L/BA.2 Omicron variant in Denmark where it became predominant over the 21K/BA.1 Omicron variant that predominated until then suggests that such epidemiological change may occur in other countries worldwide (Figure 3b, 3c).^20^ In our institute we diagnosed 16,285 SARS-CoV-2 infections between 28/11/2021 (first detection of the Omicron variant) and 02/02/2022, during which 66% of infections were identified as due to the 21K/BA.1 Omicron variant. A first study conducted in Denmark has reported a higher contagiousness with the 21L/BA.2 Omicron variant (n= 2,122 primary household patients) than with the Omicron 21K/BA.1 variant (n= 5,702 primary household patients).^20^ Secondary attack rates were 39% and 29% among households, respectively, and susceptibility to infection was reported to be significantly increased for unvaccinated (odd ratio (OR), 2.2) as well as full-vaccinated (2.5) and booster-vaccinated (3.0) people. No data to our knowledge is currently available regarding the frequency of asymptomatic and mild and severe clinical forms with this 21L/BA.2 Omicron variant.

As for the 21K/BA.1 and 21M/BA.3 Omicron variants, the origin of the 21L/BA.2 Omicron variant is currently unclear. The great number of amino acid substitutions in the spike protein and receptor binding domain of these viruses has fuelled several hypotheses that include overlooked virus evolution in people with low access to viral diagnosis and genome sequencing, in an immunocompromized chronically-infected patient, or in animals.^5^ A closest known Omicron ’s ancestor has been estimated to date back to mid-2020.^5^ Another finding is that despite the tremendous amount of genome sequences available in GISAID (7,790,928 as of 02/02/2022) we are still unable to predict the emergence, and outcome of new variants. This supports the real-time close surveillance of the emergence, spread and vanishing of SARS-CoV-2 variants through molecular and genomic surveillance. It is also worthy of interest to assess phenotypically through inoculation on permissive cells the susceptibility of emerging variants to neutralization by anti-spike antibodies elicited by prior infection or by vaccination, which is on-going in our laboratory for the 21L/BA.2 Omicron variant.

## Data Availability

Genome sequences described here were deposited in the GISAID sequence database (https://www.gisaid.org/).

## Acknowledgments

We are grateful to Ludivine Brechard, Claudia Andrieu, Emilie Burel, Elsa Prudent, Céline Gazin, and Marielle Bedotto for their technical help.

## Author contributions

Study conception and design: Philippe Colson, Didier Raoult, Jacques Fantini, and Pierre-Edouard Fournier. Materials, data and analysis tools: Philippe Colson, Jeremy Delerce, Mamadou Beye, Anthony Levasseur, Céline Boschi, Linda Houhamdi, Hervé Tissot-Dupont, Nouara Yahi, Matthieu Milllion, Jacques Fantini. Data analyses: Philippe Colson, Pierre-Edouard Fournier, Bernard La Scola, Didier Raoult, Jeremy Delerce, Mamadou Beye, Anthony Levasseur, Jacques Fantini, and Nouara Yahi. Writing of the first draft of the manuscript: Philippe Colson, Jacques Fantini, and Pierre-Edouard Fournier. All authors read, commented on, and approved the final manuscript.

## Data availability

The dataset generated and analyzed during the current study are available in the GISAID database (https://www.gisaid.org/).

## Ethics approval

This study has been approved by the ethics committee of University Hospital Institute (IHU) Méditerranée Infection (N°2022-008). Access to the patients ’ biological and registry data issued from the hospital information system was approved by the data protection committee of Assistance Publique-Hôpitaux de Marseille (APHM) and was recorded in the European General Data Protection Regulation registry under number RGPD/APHM 2019-73.

